# Stigmatization of Indigenous patients in healthcare: Co-development and validation of a measurement tool

**DOI:** 10.64898/2026.06.06.26355055

**Authors:** Marie-Claude Tremblay, Elvis Iradukunda, Christine Cassivi, Pascale Breault, Élaine Brière, Caroline Collerette, Christopher Fletcher, Jean-Sébastien Renaud, Marianne Beaulieu

## Abstract

**Introduction:** Indigenous peoples in Canada face persistent health inequities rooted in colonialism, systemic racism, discrimination and social exclusion, all of which operate with particular intensity within healthcare institutions. Despite a growing qualitative literature documenting the discrimination and stigmatisation of Indigenous people by healthcare professionals, no validated instrument existed in the Canadian context to measure the stigmatizing attitudes and behaviors of clinicians toward this population.

**Aim:** This study aimed to co-develop and validate an instrument using clinical case vignettes designed to capture the affective, cognitive, and behavioral dimensions of stigmatization of indigenous peoples.

**Method:** Following Boateng et al.’s three-phase scale development approach, a multidisciplinary team including Indigenous patient partners, researchers, clinicians, and measurement experts generated 244 items across three paired clinical vignettes addressing type 2 diabetes, chronic back pain, and depressive disorder. Each vignette was developed in two versions, one featuring an Indigenous patient (test) and one featuring a non-Indigenous patient (control), distinguished solely by name and origin. Content validity was assessed by an expert committee using a Content Validity Index. The instrument was subsequently administered to a sample of nurses and physicians from two canadian health institutions using a twelve-arm randomization design. Analyses were carried to assess the internal structure of the instrument, convergent and concurrent validity as well as internal consistency.

**Results:** Our results show that the instrument developed has good psychometric qualities, particularly in terms of internal consistency, concurrent validity and factor structure, which reflects the theoretical structure assumed. Concurrent validity of the tool with the M-PATAS scale demonstrated weak to moderate significant correlations. Developed through a participatory process centering Indigenous expertise and lived experience, this instrument constitutes a significant methodological advance in the study of racialized stigmatization in Canadian healthcare.

## 1. Introduction

In Canada as in most other countries around the world, Indigenous peoples experience significant and persistent health inequities compared to the majority population (1–4). Many of these inequities are rooted in colonialism and its aftermath, including institutional violence, intergenerational trauma, forced displacement and relocation, systemic racism, stigmatization, and social exclusion (5, 6). As a result of these systematic inequities, Indigenous populations are more likely than other groups to experience life adversity such as intergenerational poverty, social deprivation, housing issues, family violence, and chronic exposure to stress (1, 7–10). The profound impact of colonialism contributes to the stigmatization of Indigenous peoples and reinforce racist stereotypes, further subjecting them to exclusion and marginalization in a vicious cycle (7).

Racism, discrimination and stigmatization that are prevalent in Canadian society also permeate our public institutions, resulting in critical barriers to accessing the healthcare system (11–17). Despite the prevailing ethos of egalitarianism underlying the healthcare system, implicit racialization processes are persistent (13, 18) and result in First Peoples receiving “second-class treatment”, to use Allan and Smylie’s characterization (7). A sizeable body of research has documented the negative health care experiences of Indigenous peoples in Canada (12, 19–22). These studies show that Indigenous peoples’ interactions with healthcare providers are characterized by fear, intimidation, stereotyping, stigmatization and racism (12, 13, 19, 20, 23, 24). Several studies have documented healthcare professionals’ perceptions and biases toward Indigenous peoples, whom they describe as heavier cases (25), patients lacking adherence to care (25), and with pathological substance use behaviors (26, 27). Along the same lines, Harding (28) found that Indigenous patients are dehumanized and stereotyped as non-compliant and as having pathological behaviors. These stereotypes lead to certain forms of discrimination that have significant physical and emotional impacts, such as improper or abusive clinical interactions, misdiagnoses or missed diagnoses, inappropriate pain management and services avoidance, to name a few (17, 28). Similarly, Kitching et al. demonstrated that urban Indigenous populations who had experienced discrimination by a healthcare provider were more likely to have unmet health needs (29).

With a few notable exceptions (17, 28, 29), most of the research carried out in Canada on the topic of racism, discrimination and stigmatization of Indigenous patients is qualitative in essence. This research consists in interpretative, small-samples, descriptive studies exploring the experiences of Indigenous peoples in the healthcare system, or the perceptions of healthcare professionals towards these patients (12, 13, 19, 20, 24, 30–32). While this type of study is fundamental to understanding the lives and experiences of these marginalized groups from their own perspective, it does not provide a systematic view of their collective experience. In addition, from an anti-oppressive research perspective, it is necessary to shift the focus in order to better understand the dynamics of oppression, racism and discrimination perpetrated by healthcare professionals, rather than focusing once again on those who suffer injustice and inequality. Examining the attitudes, beliefs and behaviours of healthcare professionals towards Indigenous peoples appears as a valid approach to responding to the systemic racism and culturally unsafe care practices they face in the healthcare system. This project is part of an effort to deepen our knowledge of the links between attitudes, stereotypes and the stigmatizing behaviours of healthcare professionals, and to better understand the conditions under which stigmatization occurs.

## 2. Aim and specific objectives

The need for the present project was first identified by Indigenous partners involved in a previous project on cultural safety in health care (30). Community partners felt that it was necessary to empirically and objectively investigate if Indigenous patients in general were received and treated differently in the healthcare system. After a review of literature on the stigmatization of Indigenous peoples, we found that no specific instrument had been developed to measure this in the Canadian context.

This project aimed to develop and validate an instrument that uses clinical case scenarios (vignettes) to measure the stigmatization of Indigenous patients. The specific objectives of the study were:

1. to co-develop an instrument to measure stigmatization of Indigenous patients by nurses and doctors;
2. to assess the validity and reliability of the instrument developed.

Regarding the population investigated, we chose to specifically target nurses and doctors, as they represent the largest group of healthcare professionals in the healthcare system, and are most often cited in the literature as perpetrators of stigmatization. The article reports in detail the 3-phase process we followed to develop our innovative instrument namely: item development and testing, instrument development and instrument evaluation. These phases are described below.

## 3. Methods

The instrument was developed and assessed in line with the method proposed by Boateng, Neilands (33) which generally follows three phases: 1) item development; 2) scale development; 3) scale evaluation. The entire project was carried out in partnership with a multidisciplinary team comprised of measurement and evaluation experts, public health experts, Indigenous patient partners, a representative of an Indigenous organization and clinicians.

### 3.1. Phase 1: Item development

#### 3.1.1. Identification of the domain

In line with the methodology proposed by Boateng, Neilands (33), the first step involved identifying the domain (i.e. the concept measured by the instrument), generating items and assessing content validity. To this end, a literature review on the concept of stigmatization was carried out, identifying the boundaries of the field of interest and proposing an operationalized definition of the studied concept. Drawing on Link and Phelan (34), Dieujuste (35) et Nyblade, Stockton (36), we defined stigmatization as a **set of negative attitudes, negative feelings and generalized beliefs of members of a majority group towards members of a minority group, which can lead to the unfair treatment (discrimination) of members of the latter group**. Stigmatization therefore has 3 dimensions: affective, cognitive and behavioral.

The affective dimension of stigmatization represents negative or even hostile attitudes and feelings towards the Indigenous patient. It includes discomfort, unease and feelings of exacerbation towards the patient. The cognitive dimension represents negative beliefs, biases, or stereotypes that one might form against Indigenous patients. It is reflected in the attribution of negative characteristics to Indigenous patients, and often manifests itself in stereotypes regarding them. The literature review pointed not only to general stereotypes regarding Indigenous peoples, but also to more specific stereotypes of Indigenous patients. Based on this review, we identified 4 cognitive sub-dimensions:

1. prevalent stereotypes towards Indigenous peoples in society in general (e.g. alcohol and substance abuse, unreliability, dishonesty).
2. lack of autonomy, i.e the belief that Indigenous patients cannot take charge of themselves, make decisions about their care or become actively involved in their care;
3. responsibility for their condition, i.e. the belief that Indigenous patients are personally responsible for their bad health status, that they have done something wrong and deserve what they get;
4. non-compliance, i.e. the belief that Indigenous patients are less compliant with professional recommendations and prescribed treatment, and that they are thus more difficult to treat.

Finally, the affective and cognitive dimensions of stigmatization manifest themselves in a behavioral component. The behavioral dimension represents discrimination in the healthcare system, in the form of differences in treatment or clinical recommendations for Indigenous patients. It can be captured by questions about how practitioners apply clinical recommendations in given situations.

#### 3.1.2. Item generation

Three vignettes depicting clinical situations were developed by the multidisciplinary research team among whom were Indigenous partners. For each of the vignettes featuring Indigenous patients (test vignettes), identical control vignettes were developed, with the only difference being the person’s ethnocultural background, identified by the patient’s name (Indigenous or non-Indigenous consonance; i.e. “Vollant” or “Vigneault”), as well as by the city of origin (Table 1). It was decided not to explicitly identify the person as Indigenous, other than by their indigenous-sounding name and place of residence (indigenous community), in order to approximate the clinical situations in which the clinician would infer this information. The scenarios in the three vignettes address three different clinical cases: type 2 diabetes (vignette 1), chronic back pain (vignette 2) and depressive disorder (vignette 3). The themes of chronic pain and depression are inspired by vignettes used in similar studies (37, 38). These vignettes were designed for nurses regardless of training, and doctors of any specialization. The vignettes deliberately present general situations, so that most participating nurses and doctors, whatever their specialty, could understand and respond to them. Following the development of the vignettes, items related to each of the dimensions of stigmatization (affective, cognitive and behavioral) were generated by the research team (n=244 in total for the 3 vignettes). Each of these items is rated on a 6-point Likert scale with answers ranging from strongly agree (1) to strongly disagree (6). This scale deliberately avoids a neutral point and forces participants to lean in one direction or the other. Examples of items related to the affective dimension of stigmatization are: “I would prefer to avoid caring for Mr. Vollant”, “I am uncomfortable caring for patients like Mr. Vollant”, “I am empathetic towards Mr. Vollant” (reverse coded item). Items relating to the cognitive dimension of stigmatization play on the various stereotypes and prejudices towards Indigenous patients identified in the literature, and include, for example: “It is the health professional who must decide on the best treatment options for Mr. Vollant”, “Mr. Vollant’s state of health results from his poor lifestyle habits”, “Mr. Vollant seems to be a cooperative patient” (reversed coded item). As for the behavioral dimension of stigmatization, its assessment is based mainly on the assessment of the severity of the patient’s symptoms (i.e. “on the basis of the information available, please assess the severity of the symptoms Mr. Vollant is experiencing today”), and questions designed to examine the prioritization of recommendations that the professional would make (variable from one situation to another).

**Table 1.** Clinical vignette on Type 2 diabetes.

| Indigenous patient (test vignette) | Non-Indigenous patient (control vignette) |
| --- | --- |
| Patient Description: Mr. Vollant<br>- 45-year-old, overweight patient<br>- Seasonal worker<br>- Lives in the Indigenous community of Pessamit (Québec)<br>- Married, father of three<br>Mr. Vollant was diagnosed with type 2 diabetes two years ago, while hospitalized for cellulitis. Since then, Mr. Vollant has made frequent visits to the hospital for hyperglycemia, which he has been unable to control. His most recent test showed an elevated glycated hemoglobin (HbA1c). Today, he presents again with hyperglycemia. He is also showing signs of severe dehydration and drowsiness. Mr. Vollant says he does his best to control his blood sugar, eat well and exercise frequently, but finds it difficult to reach treatment targets. | Patient Description: Mr. Vigneault<br>- 45-year-old, overweight patient<br>- Seasonal worker<br>- Lives in Baie-Comeau (Québec)<br>- Married, father of three<br>Mr. Vigneault was diagnosed with type 2 diabetes two years ago, while hospitalized for cellulitis. Since then, Mr. Vigneault has made frequent visits to the hospital for hyperglycemia, which he has been unable to control. His most recent test showed an elevated glycated hemoglobin (HbA1c). Today, he presents again with hyperglycemia. He is also showing signs of severe dehydration and drowsiness. Mr. Vigneault says he does his best to control his blood sugar, eat well and exercise frequently, but finds it difficult to reach treatment targets. |

*3.1.3. Content validity*

#### 3.1.3. Content validity

To assess the content validity of the instrument, a committee of 9 Indigenous health experts who were not part of the research team was set up. The committee was composed of researchers with expertise in public health, Indigenous health or stigmatization, as well as Indigenous nurses and physicians, and health professionals working with this clientele. Using a 5-point Likert scale, the expert committee was asked to assess the clarity (1-not clear at all, 5-very clear) and relevance (1-not relevant at all, 5-very relevant) of each of the items in the three vignettes in relation to the conceptual definition of stigmatization and its dimensions (33). A Content validity index was calculated for each item (number of experts rating 4 or more)/(total number of experts). Items with a CVI (39) of more than 0.85 in terms of clarity and relevance were selected for inclusion in the initial version of the instrument, for a total of 96 items. Of these, 29 items were common to all three vignettes, while 9 vignette-specific questions were designed to assess the behavioral dimension of stigmatization. Examples of the items selected are shown in Table 2.

**Table 2.**
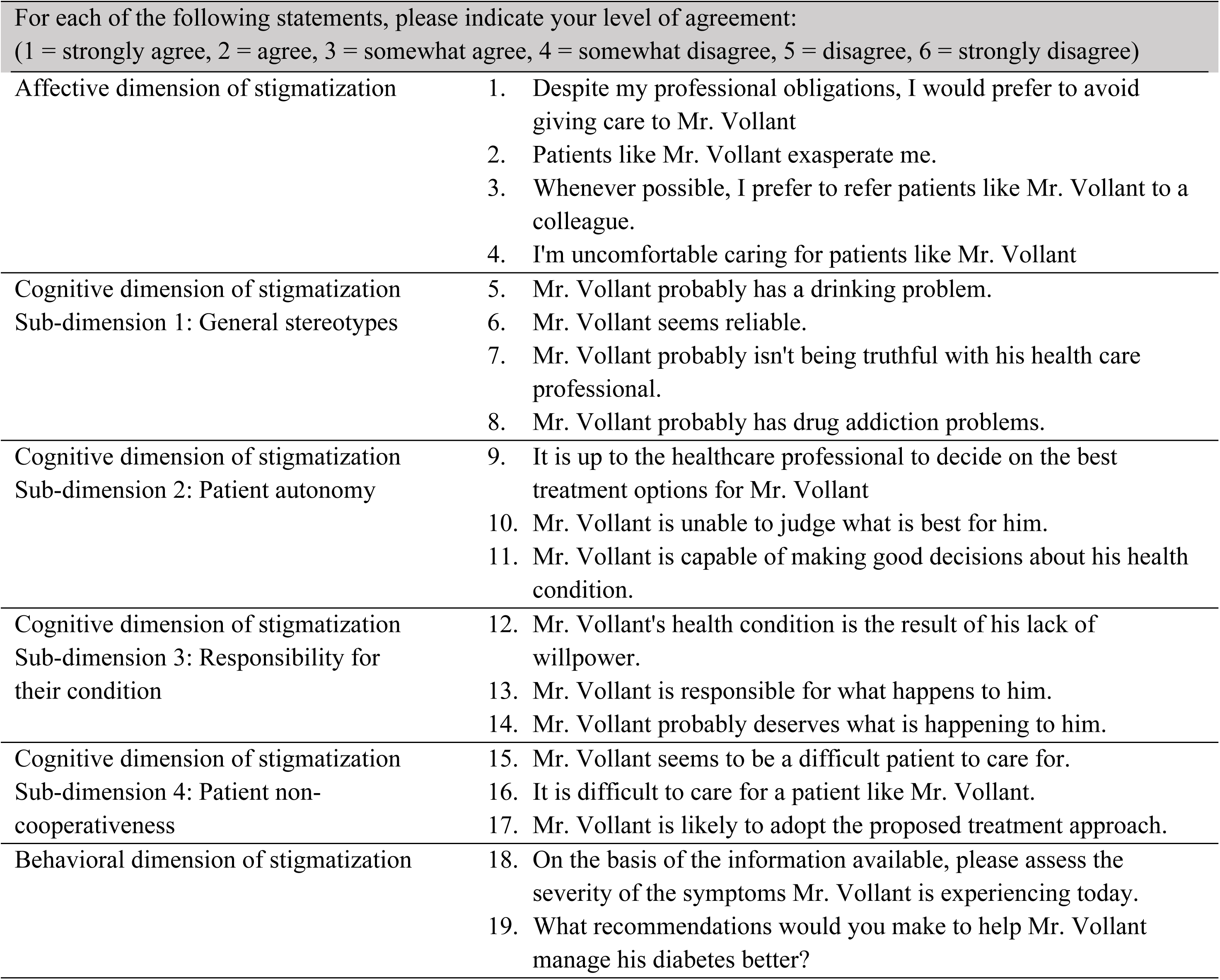
Examples of selected items.

| For each of the following statements, please indicate your level of agreement:<br>(1 = strongly agree, 2 = agree, 3 = somewhat agree, 4 = somewhat disagree, 5 = disagree, 6 = strongly disagree) |  |
| --- | --- |
| Affective dimension of stigmatization | 1. Despite my professional obligations, I would prefer to avoid giving care to Mr. Vollant<br>2. Patients like Mr. Vollant exasperate me.<br>3. Whenever possible, I prefer to refer patients like Mr. Vollant to a colleague.<br>4. I'm uncomfortable caring for patients like Mr. Vollant |
| Cognitive dimension of stigmatization<br>Sub-dimension 1: General stereotypes | 5. Mr. Vollant probably has a drinking problem.<br>6. Mr. Vollant seems reliable.<br>7. Mr. Vollant probably isn't being truthful with his health care professional.<br>8. Mr. Vollant probably has drug addiction problems. |
| Cognitive dimension of stigmatization<br>Sub-dimension 2: Patient autonomy | 9. It is up to the healthcare professional to decide on the best treatment options for Mr. Vollant<br>10. Mr. Vollant is unable to judge what is best for him.<br>11. Mr. Vollant is capable of making good decisions about his health condition. |
| Cognitive dimension of stigmatization<br>Sub-dimension 3: Responsibility for their condition | 12. Mr. Vollant's health condition is the result of his lack of willpower.<br>13. Mr. Vollant is responsible for what happens to him.<br>14. Mr. Vollant probably deserves what is happening to him. |
| Cognitive dimension of stigmatization<br>Sub-dimension 4: Patient non-cooperativeness | 15. Mr. Vollant seems to be a difficult patient to care for.<br>16. It is difficult to care for a patient like Mr. Vollant.<br>17. Mr. Vollant is likely to adopt the proposed treatment approach. |
| Behavioral dimension of stigmatization | 18. On the basis of the information available, please assess the severity of the symptoms Mr. Vollant is experiencing today.<br>19. What recommendations would you make to help Mr. Vollant manage his diabetes better? |

### 3.2. Phase 2: Scale development

#### 3.2.1. Pre-testing of questions

To ensure that the terms used in the clinical vignettes and instrument items were relevant and consistent in describing and measuring the phenomenon under study, for the target population, two waves of item testing were carried out with a convenience sample of the instrument’s target population: three nurses and four physicians (n=7; wave 1) and three nurses and one physician (n=4; wave 2). Pre-test participants were asked to read the vignettes, answer the items and then respond to qualitative interview questions about the questionnaire and their understanding of the items. This phase was used to improve certain aspects of the instrument, the formatting and flow of the questions. This testing also led to the reformulation of certain elements deemed unclear or confusing.

#### 3.2.2. Administration of the instrument

The final stages of the instrument’s development involved administering the questionnaire to a larger sample of nurses and physicians. A convenience sample was recruited on a voluntary basis from physicians and nurses (n=215) from the Centre intégré de santé et services sociaux (CIUSSS) de la Capitale-Nationale and from the Centre hospitalier universitaire de Québec-Université Laval (CHU-UL). Data were collected from September to November 2020, as well as from January to March 2021. Invitations to participate in the study were sent to various departments in these two organizations (Family Medecine Groups, Public Health, Intellectual Disability Services, Autism Spectrum Disorder Services, Mental Health and Addiction Program, Geriatric Services, Multi-disciplinary Services, External clinic, Anesthesiology, Surgery, Medical Imaging, General Medicine, Emergency Medicine, Obstetrics and Gynecology, Pediatrics). Sample selection criteria included: 1) working as a registered nurse or physician in Quebec (Canada); 2) being able to answer an online questionnaire written in French.

At this point, the questionnaire included a pair of randomly selected vignettes, comprising one vignette in the test version (29 items + 2 to 5 multiple-choice questions) and one vignette in the control version (29 items + 2 to 5 multiple-choice questions). Survey administration used a 12-arm randomization strategy, allowing for all crossovers between pairs of 3 clinical vignettes, treatments (test vignette vs. control vignette) as well as the order of passage of the test vs. control vignette (first vs. second position). The randomization groups are presented in Table 3. The questionnaire was imported into an online platform (Qualtrics) and the online survey was completed independently by participants.

**Table 3.**
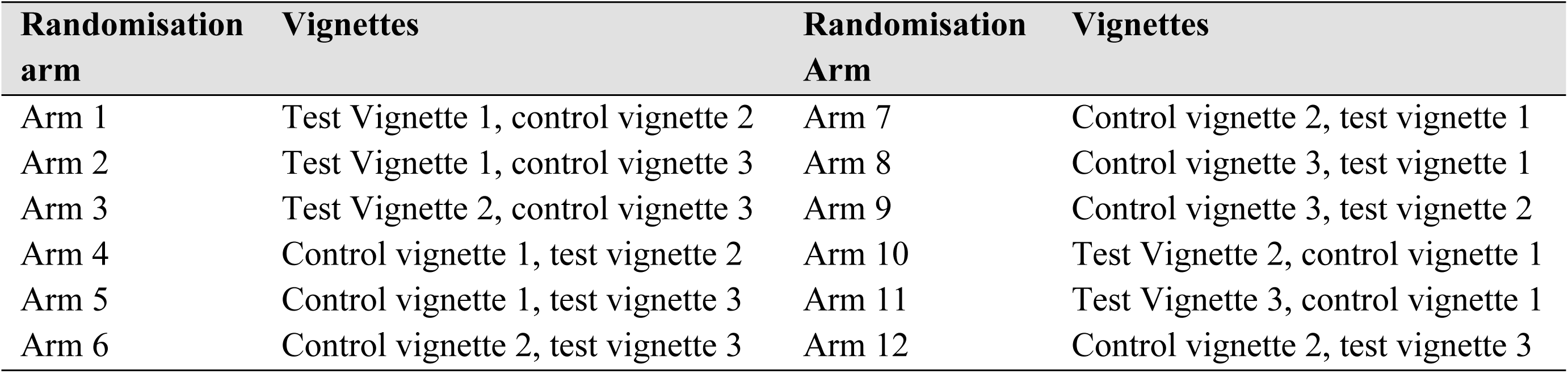
Randomized groups.

The questionnaire also contained socio-demographic questions (age, gender, belonging to a visible minority or Indigenous group) and questions related to practice characteristics (region of practice, professional discipline, years of experience, type of healthcare organization, plus frequency of contact with Indigenous people). The questionnaire also included the M-PATAS scale, a validated instrument designed to measure modern prejudice towards Indigenous people (40, 41). This scale, which was used to assess convergent validity of our instrument, contains 14 items measured in a 5-point Likert-type response format (1-strongly disagree; 5-strongly agree). The possible range of scale scores varies from 14 to 70, with greater scores interpreted as greater levels of prejudicial attitudes towards Indigenous people.

All participants provided informed consent for their participation in the study. As the questionnaire was anonymous and electronic, submitting it was considered an expression of consent. Due to the sensitive nature of the topic studied and the social desirability biases it may entail, the actual aims of the study were only revealed at the end of the survey, with the possibility for participants to withdraw their data from the project. This strategy and the study were approved by the CIUSSS and the CHU-UL ethics boards (project no MP13-2019-1793; 2019-1793_SPPL; 2021-4982).

#### 3.2.3. Item reduction and extraction of factors

Iteration of principal component analysis and classical item analysis were used to assess the internal structure of the instrument. The aim was to verify whether the factor structure of the instrument reflects the assumed theoretical structure. In order to determine the structure of the tool, we had to determine a common structure between the responses to Indigenous vignettes and non-indigenous vignettes. We used all the responses (Indigenous and non-indigenous vignettes) in order to simplify the tool and reduce the number of items. Items with factor loading greater than .400 were retained [49], while items displaying cross-loading (loadings on two or more factors concurrently) were removed.

### 3.3. Scale evaluation

#### 3.3.1. Effects of the design

Analyses were conducted to assess potential biases induced by the study instrument and design. To this end, we first checked that the order of treatment (test vignette vs. control vignette) and the choice and order of clinical vignettes did not affect the results, made possible by the twelve-arm randomization. This analysis was carried out using a linear mixed model evaluating the effects of the vignette pair chosen, the order of vignettes, the order of treatment, the effect of vignette position, the effect of clinical vignette, the effect of treatment and the interaction between vignette and treatment on instrument dimension scores and the total score.

#### 3.3.2. Tests of validity

We assessed convergent validity and concurrent validity (42–44). Convergent validity represents the ability of an instrument to produce results similar to another instrument measuring the same construct (33, 42, 44). For this study, the instrument scores (sub-dimensions and item scores) were examined along the M-PATAS scale (Modern Prejudiced Attitudes toward Aboriginals Scale) (40, 45), an instrument including 14 items that measures a similar concept (prejudice against Indigenous) on 5 point Likert items. For this purpose, Spearman’s Rho were used. Correlations were interpreted as follows: 1.00 = perfect; 0.99-0.70 = strong; 0.40-0.69 = moderate; 0.01-0.39 = weak; 0.00 = none, in line with the work of numerous authors (46–48). To assess concurrent validity, we evaluated correlations between instrument’s dimensions, and overall score as well as score for behavioral dimensions (specific clinical recommendations).

#### 3.3.3. Tests of reliability (internal consistency)

Cronbach’s Alphas were used to assess internal consistency. They were calculated for each dimension, as well as for all items, on the scores for each of the 6 scenarios, and then on the difference between the Indigenous and non-indigenous vignettes for each of the 3 pairs of vignettes separately. The aggregated scores were obtained by subtracting the Indigenous vignette scores from the non-indigenous vignette scores.

## 4. Results

### 4.1 Administration of the instrument

A total of 219 participants completed the survey. Table 4 provides a portrait of the participants.

**Table 4.** Phase 1 survey participants.

| Variables | n (%) or M±s.d<br>(total =219) |
| --- | --- |
| Identifies as a woman | 192 (87.67) |
| Age | 39.75 ± 9.73 |
| Identifies as an Indigenous | 4 (1.83) |
| Clinical experience (number of years) | 14.18 ± 9.75 |
| Recruited at the CIUSS | 176 (80.37) |
| Recruited at the CRCHU | 43 (19.63) |
| Physicians | 63 (28.77) |
| • Family physicians and emergency physicians | 45 (20.55) |
| Nurses | 149 (68.04) |
| • Nurse clinicians | 92 (42.01) |
| Hospital or family medicine group (care setting) | 135 (61.65) |
| Report having contact with Indigenous patients a few times a year | 96 (43.84) |
| Reports having had no training on indigenous cultures/realities | 120 54.79) |

### 4.1. Item reduction and extraction of factors

A principal component analysis (Varimax rotation with Kaiser Normalization) showed 6 components for each set of data, that basically reflected the dimensions of the tool. When comparing the structures of the two sets of data (Indigenous vignettes vs. non-indigenous vignettes), we retained the items common to both (using all responses), which allowed us to reduce the number of Likert items from 29 to 21 (Table 5).

**Table 5.**
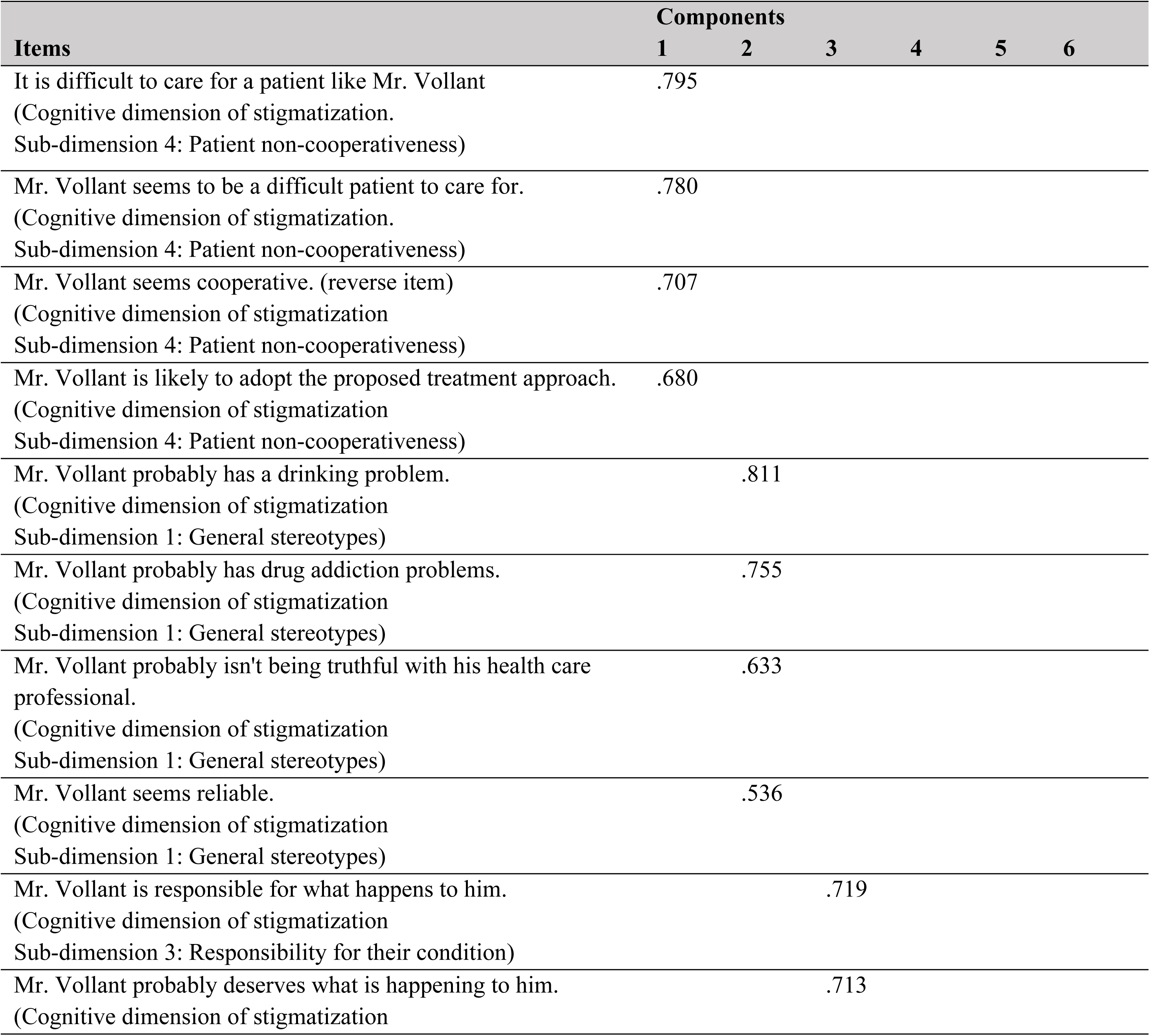
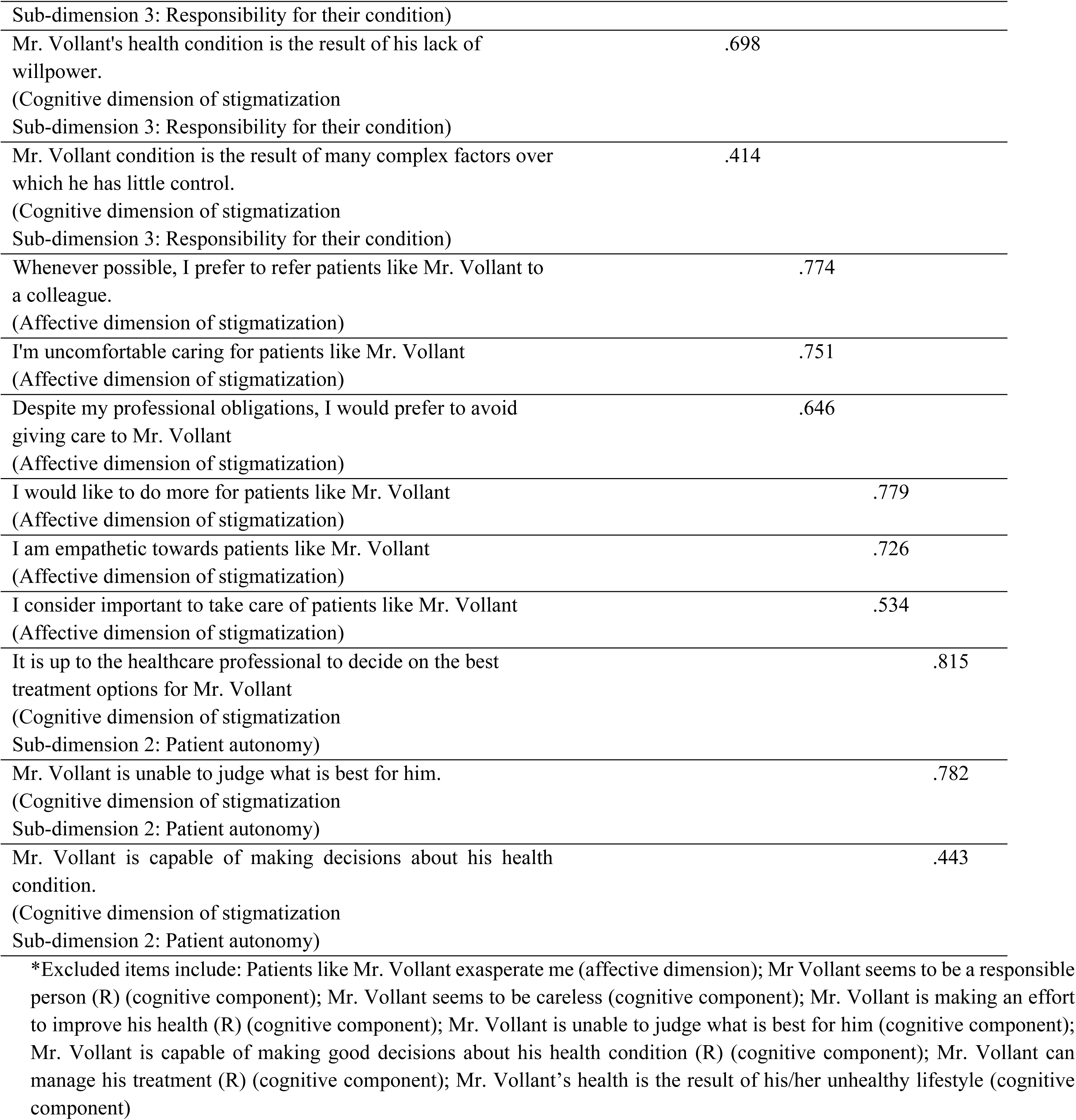
selected items. *Excluded items include: Patients like Mr. Vollant exasperate me (affective dimension); Mr Vollant seems to be a responsible person (R) (cognitive component); Mr. Vollant seems to be careless (cognitive component); Mr. Vollant is making an effort to improve his health (R) (cognitive component); Mr. Vollant is unable to judge what is best for him (cognitive component); Mr. Vollant is capable of making good decisions about his health condition (R) (cognitive component); Mr. Vollant can manage his treatment (R) (cognitive component); Mr. Vollant’s health is the result of his/her unhealthy lifestyle (cognitive component)

### 4.2. Effects of the design

Analysis showed that the median cognitive dimension score under the “General stereotypes” sub-dimension for participants who received vignette 1 (type 2 diabetes situation), whether Indigenous or non-Indigenous, is high compared with the scores of participants who received other clinical vignettes. As a function of treatment, this score is almost identical for the Indigenous and non-Indigenous vignette (Figure 1). Figures 2 and 3 show no significant differences in mean and weighted mean scores between clinical vignettes 1, 2, and 3 (i.e. type 2 diabetes, back pain and depressive disorder) and in terms of treatment (Indigenous and non-Indigenous).

**Figure 1.**
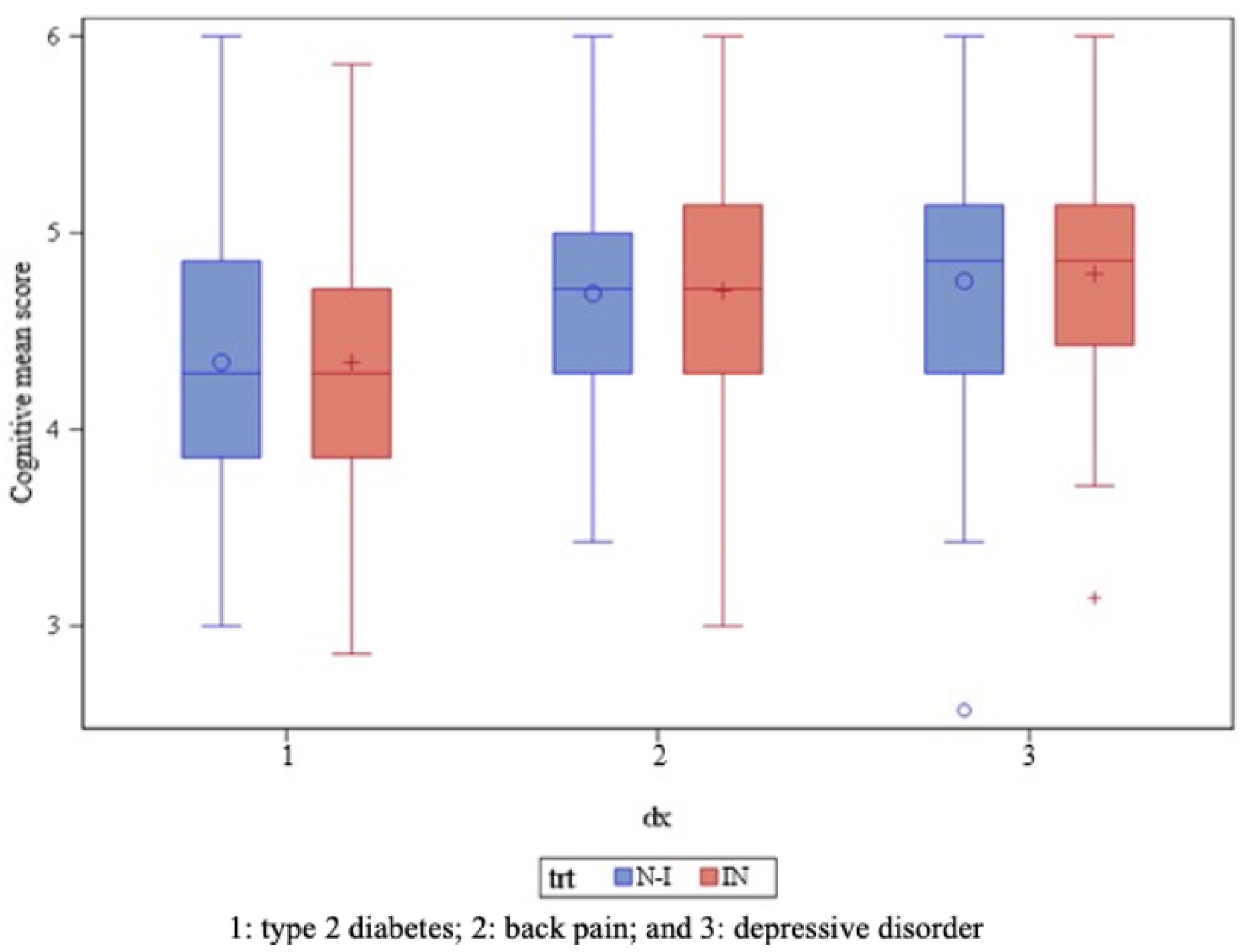
Cognitive scores under General stereotypes sub-dimension according to clinical vignettes and treatment (non-Indigenous vs Indigenous)

**Figure 2.**
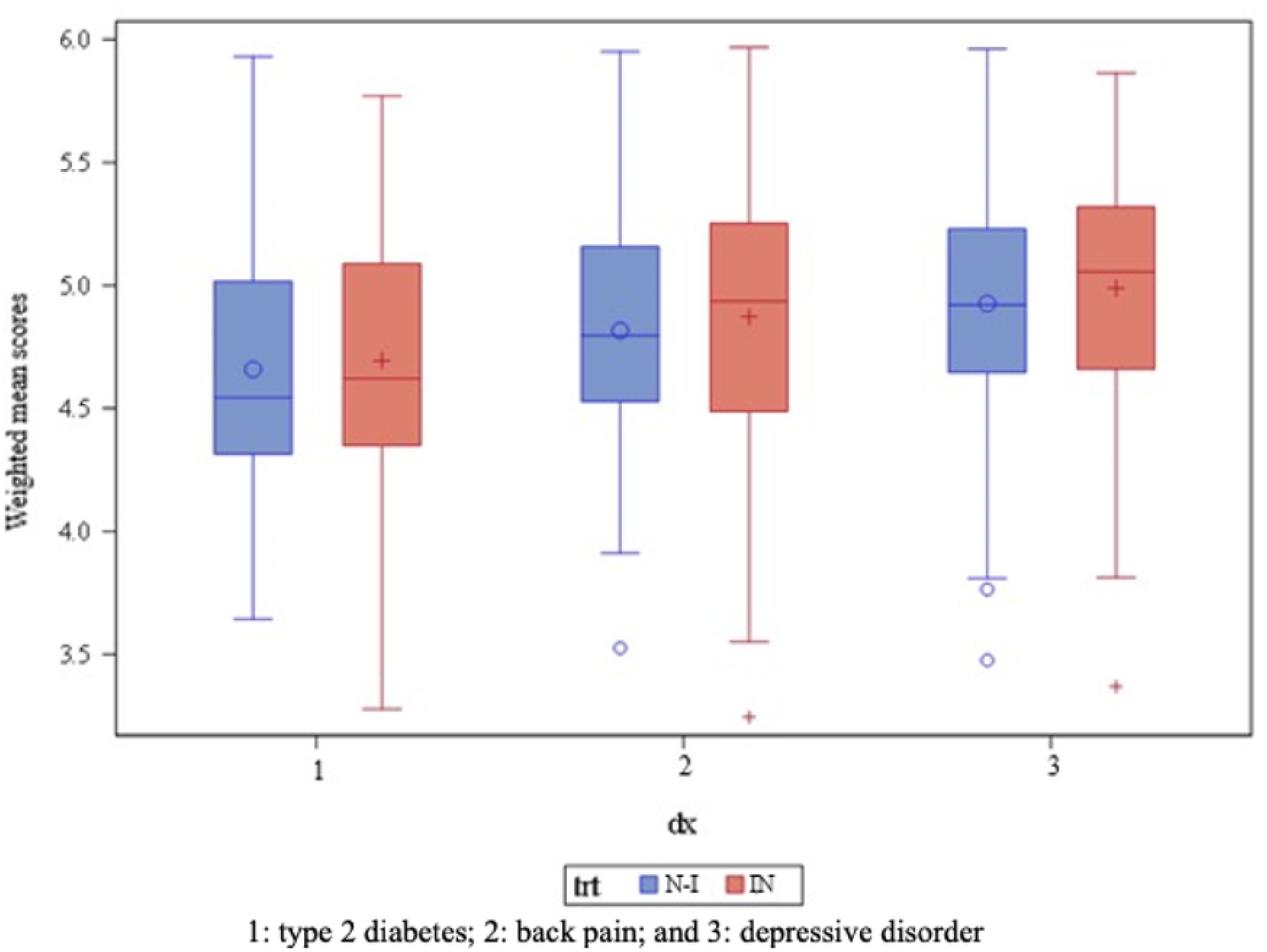
Weighted mean scores by clinical vignette and treatment (non-Indigenous vs Indigenous)

**Figure 3.**
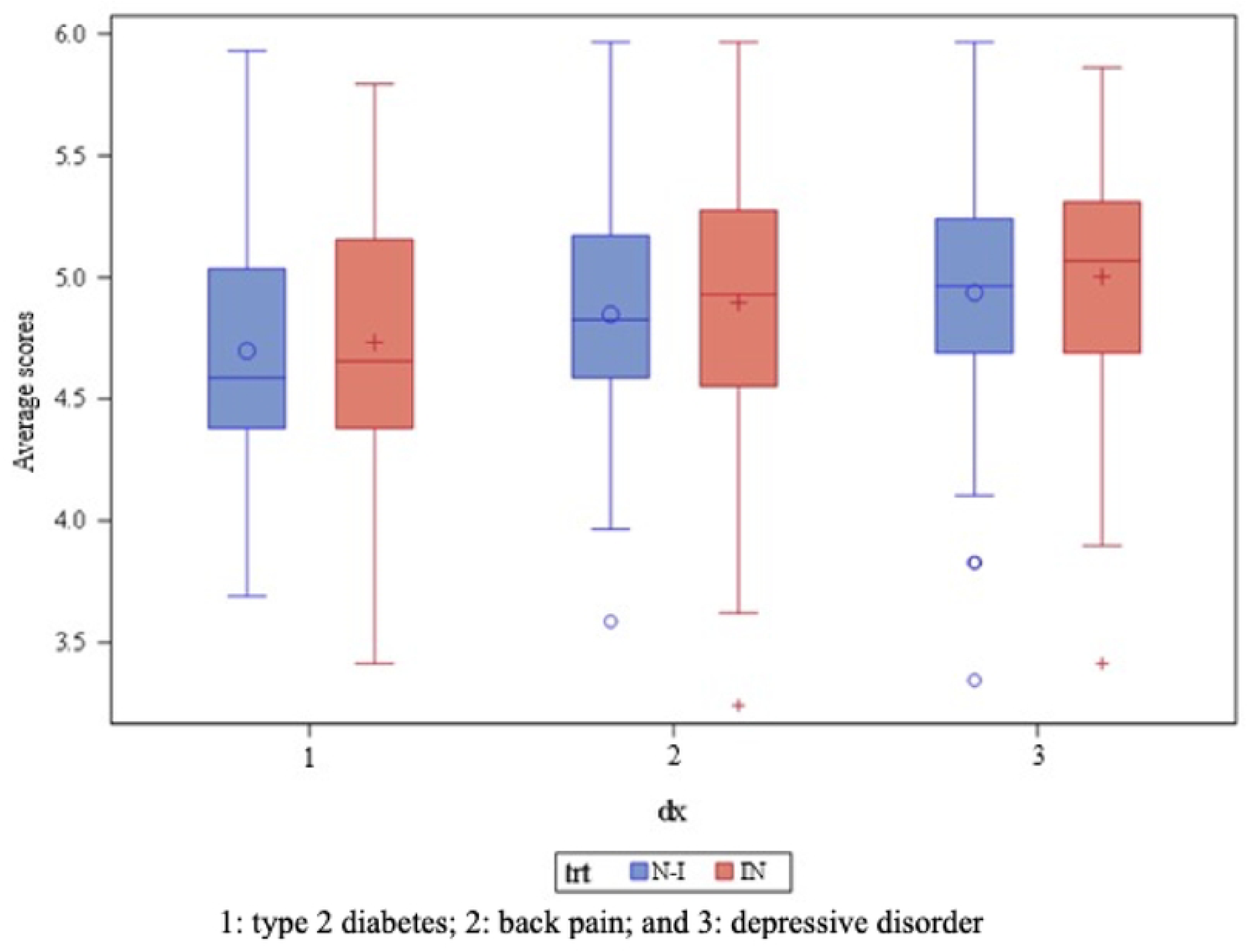
Average scores by clinical vignette and treatment (non-Indigenous vs Indigenous)

We observed the existence of the treatment order effect on the affective dimension scores, with a p-value of 0.0004. In addition, the effects of treatment order (p-value = 0.0298) and clinical vignettes (p-value < 0.0001) on the cognitive sub-dimension prevalent stereotypes scores are statistically significant. For the cognitive sub-dimension patient autonomy, treatment showed a significant effect on scores with a p-value of 0.0127. Scores under cognitive sub-dimension patient responsibility could be influenced by treatment order (p-value = 0.0067 and 0.0019) as well as clinical vignette order (p-value < 0.0001 and < 0.0001). Weighted mean scores were calculated for all dimensions. Importantly, treatment order (p-value = 0.0008) and clinical vignette order (p-value < 0.0001) had a statistically significant effect on these weighted mean scores (Table 3). With the adjusted scores, only the treatment (Indigenous or non-Indigenous) had a significant effect on the cognitive sub-component patient autonomy scores with a p-value < 0.0092. This impact did not vary according to the order of treatment or vignette (Table 6).

**Table 6.** Effect test with initial scores (Type 3 Tests of Fixed Effects)

| Dimensions and subdimensions | Effect | Num DF | Den DF | F Value | Pr > F |
| --- | --- | --- | --- | --- | --- |
| Affective dimension | the order of treatment | 1 | 213 | 12.74 | 0.0004 |
|  | the order of treatment | 1 | 213 | 4.79 | 0.0298 |
| Cognitive subdimension: Prevalent stereotypes | the order of clinical vignettes | 2 | 213 | 15.18 | <.0001 |
| Cognitive subdimension : Patient autonomy | The order of treatment | 1 | 213 | 6.31 | 0.0127 |
| Cognitive subdimension: Patient responsibility | the order of treatment | 1 | 213 | 7.50 | 0.0067 |
|  | the order of clinical vignettes | 2 | 213 | 58.99 | <.0001 |
| Cognitive subdimension: Patient non-cooperativeness | the order of treatment | 1 | 213 | 9.86 | 0.0019 |
|  | the order of clinical vignettes | 2 | 213 | 11.02 | <.0001 |
| Weighted mean | the order of treatment | 1 | 213 | 11.57 | 0.0008 |
|  | the order of clinical vignettes | 2 | 213 | 12.67 |  |
| Global mean | the order of treatment | 1 | 213 | 12.02 | 0.0006 |
|  | the order of clinical vignettes | 2 | 213 | 11.42 |  |

### 4.3. Tests of validity

Convergent validity was assessed along the M-PATAS scale using Spearman’s Rho. Since a high score on the M-PATAS scale indicates a high level of stigmatization, and a high score on our instrument indicates a low level of stigmatization, we would expect a negative correlation. We did observe a weak negative correlation, statistically significant at the 5% threshold, between scores on the M-PATAS scale and corrected scores on the affective component (Rho= −0.14, p=0.03), the prevalent stereotypes sub-dimension (Rho= −0.20, p=0.003), patient autonomy sub-dimension (Rho= −0.32, p < 0.0001), patient responsibility sub-dimension (Rho= −0.26, p < 0.0001), patient non-cooperation sub-dimension (Rho= −0.14, p=0.04), weighted mean scores (Rho= −0.31, p < 0.0001), and mean scores (Rho = −0.30, p < 0.0001) (Table 7).

**Table 7:** Test of correlation of Indigenous corrected scores with M-PATAS scores.

| Components | Rho | P-value |
| --- | --- | --- |
| Affective component | -0.14369 | 0.0336 |
| Cognitive subdimension : Prevalent stereotypes | -0.19732 | 0.0034 |
| Cognitive subdimension : Patient autonomy | -0.32126 | <0.0001 |
| Cognitive subdimension : Patient responsibility | -0.26277 | <0.0001 |
| Cognitive subdimension : Patient non-cooperativeness | -0.14093 | 0.0372 |
| Mean scores | -0.30204 | <0.0001 |
| Weighted mean score | -0.30723 | <0.0001 |

Concurrent validity was assessed using correlations between instrument’s dimensions and overall score and behavioral dimensions (specific clinical recommendations). For participants who received vignette 1 in Indigenous version, we observed a correlation between the various scores and specific clinical recommendations. For example, a weak negative correlation was observed between the M-PATAS score and symptom severity (r = −0.307, p = 0.0109), as well as between the certain component scores and specific clinical recommendations (Table 8).

**Table 8:** vignette 1 on type 2 diabetes – Indigenous version Severe: perception of severity of symptoms, **Hydratation:** Advice to stay well hydrated, **Mesure:** Advice to measure blood sugar levels more regularly, **Diet:** Advice to pay attention to your diet and be more physically active, **Alcohol:** Advice to limit alcohol and other drug consumption, **Review TRT:** Advice to review your treatment plan with your physician

| Variables | severe | hydratation | Mesure | Diet and PA | Alcohol | Review TRT |
| --- | --- | --- | --- | --- | --- | --- |
| **M-Patas** | **-0.307**<br>**0.0109** | -0.09212<br>0.4550 | -0.05459<br>0.6584 | -0.14106<br>0.2512 | -0.11845<br>0.3360 | 0.23756<br>0.0511 |
| **Affective component** | -0.12754<br>0.3000 | 0.14214<br>0.2476 | 0.14487<br>0.2385 | -0.09461<br>0.4428 | **-0.25128**<br>**0.0387** | 0.08344<br>0.4987 |
| **Cognitive subdimension : prevalent stereotypes** | 0.07617<br>0.5370 | -0.03348<br>0.7863 | **0.25615**<br>**0.0350** | 0.04906<br>0.6912 | **0.29248**<br>**0.0155** | -0.15439<br>0.2087 |
| **Cognitive subdimension : Patient autonomy** | 0.10425<br>0.3975 | 0.19902<br>0.1037 | 0.13964<br>0.2561 | **0.25464**<br>**0.0361** | -0.09392<br>0.4462 | **-0.29661**<br>**0.0140** |
| **Cognitive subdimension : Patient responsibility** | 0.12033<br>0.3284 | **0.26079**<br>**0.0317** | 0.02438<br>0.8435 | **0.35352**<br>**0.0031** | 0.05871<br>0.6344 | **-0.40679**<br>**0.0006** |
| **Weighted mean score** | 0.08271<br>0.5025 | 0.14103<br>0.2513 | 0.19339<br>0.1141 | **0.24634**<br>**0.0429** | -0.01936<br>0.8755 | **-0.26292**<br>**0.0303** |
| **Mean score** | 0.08194<br>0.5065 | 0.12155<br>0.3235 | 0.21171<br>0.0831 | 0.22390<br>0.0664 | -0.00695<br>0.9552 | **-0.24342**<br>**0.0455** |

For those who received the second vignette (lower back pain), weak negative correlations were also found between certain component scores and clinical recommendations such as the prescription of an opioid analgesic (r= −0.37943 with a p-value of 0.0009) (Table 9). For those who received the third vignette (depression), we observed a correlation of different scores with some specific clinical recommendations and perceptions, such as the perception that the patient condition justified a sick leave. (Table 10)

**Table 9:** vignette 2 on lower back pain – Indigenous version Add test: Perform additional tests (e.g. blood test, CT scan) **Continue TRT:** Continue initial treatment and refer to a specialist **Opioid:** Prescribe/apply for prescription of opioid analgesic, **Family:** Ask more questions about family situation and living environment

| Variables | Add test | Continue TRT | Opioid | Family |
| --- | --- | --- | --- | --- |
| **Affective component** | -0.16187<br>(0.1682) | 0.13886<br>(0.2380) | **-0.37943**<br>**(0.0009)** | 0.22213<br>(0.0571) |
| **Cognitive subdimension :**<br>**prevalent stereotypes** | -0.09335<br>(0.4289) | 0.24955<br>(0.0320) | **-0.43632**<br>**(0.0001)** | 0.15234<br>(0.1951) |
| **Cognitive subdimension :**<br>**Patient non-**<br>**cooperativeness** | **-0.29747**<br>**(0.0101)** | 0.25180<br>(0.0304) | -0.34164<br>(0.0029) | 0.31746<br>(0.0058) |
| **Weighted mean score** | -0.21709<br>(0.0632) | 0.28879<br>(0.0126) | -0.39514<br>(0.0005) | 0.21494<br>(0.0659) |
| **Mean score** | -0.21445<br>(0.0665) | **0.28499**<br>**(0.0139)** | **-0.41037**<br>**(0.0003)** | 0.22417<br>(0.0548) |

**Table 10:** vignette 3 on depressive disorder – Indigenous version, Sick leave: perception that the patient condition justified a sick leave**, Abuse disorder:** clinical impression of a substance abuse disorder**, Youth protection:** Likelihood to file a report with the youth protection authorities

| Variables | Sick leave | Abuse disorder | Youth protection |
| --- | --- | --- | --- |
| **Cognitive subdimension :**<br>**prevalent stereotypes** | 0.32787<br>(0.0036) | **0.32344**<br>**(0.0041)** | -0.26618<br>(0.0193) |
| **Cognitive subdimension :**<br>**Patient autonomy** | **0.26524**<br>**(0.0197)** | -0.07021<br>(0.5440) | -0.22543<br>(0.0487) |
| **Weighted mean score** | **0.33004**<br>**(0.0034)** | 0.18640<br>0.1045 | **-0.24519**<br>**(0.0316)** |
| **Mean Score** | **0.33393**<br>**0.0030** | 0.20013<br>0.0810 | **-0.24914**<br>**0.0289** |

#### 4.3.1. Tests of reliability (internal consistency)

Overall, internal consistency proved to be very good, as shown by most situations where Cronbach’s Alpha coefficients exceed 0.75. It is moderate for scenarios with a Cronbach’s Alpha coefficient between 0.50 and 0.75, while it is poor for scenarios where the Cronbach’s Alpha coefficient is below 50%, as is the case for the stigma scores of the second vignette (low back pain), affective component (Cronbach’s Alpha=0.35) (Table 11).

**Table 11:** Internal consistency of all clinical vignettes on each dimension using Cronbach’s Alpha.

| Clinical vignettes | Component<br>(Sub-component) | Alpha_indigenous | Alpha_non-indigenous | Alpha_Stigmatization |
| --- | --- | --- | --- | --- |
| 1 | Affective | 0.738055 | 0.691902 | 0.688856 |
|  | Cognitive<br>(Prevalent<br>stereotypes) | 0.816617 | 0.879611 | 0.853251 |
|  | Cognitive<br>(autonomy) | 0.791503 | 0.762051 | 0.677888 |
|  | Cognitive<br>(responsibility) | 0.768316 | 0.686806 | 0.711594 |
|  | Cognitive<br>(non-cooperativeness) | 0.818404 | 0.879453 | 0.748015 |
|  | All items | 0.911630 | 0.908672 | 0.880944 |
| 2 | Affective | 0.819818 | 0.710171 | 0.351237 |
|  | Cognitive<br>(Prevalent<br>stereotypes) | 0.849623 | 0.819881 | 0.701771 |
|  | Cognitive<br>(autonomy) | 0.784740 | 0.829169 | 0.562528 |
|  | Cognitive<br>(responsibility) | 0.695353 | 0.689316 | 0.629133 |
|  | Cognitive (non-cooperativeness) | 0.868547 | 0.728467 | 0.726216 |
|  | All items | 0.932337 | 0.895629 | 0.747063 |
|  | Affective | 0.724850 | 0.812395 | 0.599141 |
| 3 | Cognitive<br>(Prevalent<br>stereotypes) | 0.844843 | 0.847533 | 0.734815 |
|  | Cognitive<br>(autonomy) | 0.780811 | 0.780740 | 0.646829 |
|  | Cognitive<br>(responsibility) | 0.646529 | 0.615826 | 0.648059 |
|  | Cognitive (non-cooperativeness) | 0.819975 | 0.836126 | 0.769667 |
|  | All items | 0.902022 | 0.912372 | 0.846997 |

## 5. Discussion

Based on a collaborative approach integrating different types of expertises (patient, clinical and scientific), this study aimed to co-develop and validate an instrument that uses clinical case scenarios (vignettes) to measure the stigmatization of Indigenous patients by healthcare professionals. To this end, we developed three clinical scenarios on type 2 diabetes, chronic back pain and depressive disorder. Each was used to create two almost identical vignettes, the only difference being one presenting an indigenous patient and the other, a non-indigenous patient. Our results show that the instrument developed has good psychometric qualities, particularly in terms of concurrent validity and factor structure. Indeed, the overall factor structure of the instrument reflects the theoretical structure originally assumed and the factors demonstrate good internal consistency. Concurrent validity of the tool with the M-PATAS scale demonstrated weak to moderate significant correlations between the M-PATAS score and scores for each of the dimensions, as well as with the total score. This can be explained by the fact that these two instruments measure similar—yet ultimately different—concepts (stigmatization in healthcare context vs. general stereotypes). Score analysis revealed statistically significant differences between vignettes for some items, highlighting increased stigmatization of the Indigenous patient in comparison with the non-indigenous patient. In particular, dimensions related to discomfort in treating an Indigenous patient and perception of their autonomy showed high sensitivity to cultural bias. However, our study showed that vignettes may not be equivalent to one another, and the order of administration of the vignettes might influence responses. Future work will involve re-testing and adapting the instrument, for instance by including only one vignette. In this case, the first vignette (Type 2 diabetes) would be the better choice because several items associated with this vignette show significant effect size in the anticipated direction.

Our research has several theoretical and clinical implications. From a theoretical point of view, the majority of studies on stigmatization of Indigenous people in healthcare institutions are qualitative and carried out on small samples (12, 13, 19, 20, 24, 30–32). Our study enriches the literature by developing and validating a quantitative tool that provides an overall picture of the different dimensions (affective, cognitive and behavioral) of stigmatization. By contextualizing stigmatization from a clinical point of view and according to its different dimensions, our study deepens our understanding of how stigmatization operates within healthcare environments and may affect quality of clinical care and recommendations. Furthermore, by shedding light on how stigmatization manifests in a clinical situation, this instrument may enable clinicians to recognize and address their own biases, ultimately improving cultural safety and quality of care for Indigenous patients.

## Strengths and limitations

Our studies have several strengths. Indeed, the instrument is innovative, providing an objective measure of stigma towards Indigenous patients. The instrument’s relevance is strengthened by the collaboration with a multidisciplinary team, which included indigenous partners, healthcare professionals and measurement and evaluation experts. This participatory methodology not only reinforces the instrument’s cultural sensitivity but also ensures that it addresses real prejudices and barriers encountered by Indigenous patients. By placing the lived experience of Indigenous partners at the heart of the research process, this study provides a coherent, contextually grounded instrument that aligns with the clinical reality and unique needs of Indigenous people receiving care, making it a valuable asset for promoting culturally safe healthcare practices.

Despite its strengths, this study has some limitations. First, the instrument was developed and tested with nurses and physicians, which may limit its relevance and applicability to other healthcare professionals who also interact with Indigenous patients, such as social workers and psychologists. Also, the sample for this study was relatively small (n=215). Studies with a larger sample are needed to validate the generalizability and reliability of this instrument across different healthcare settings. Additionally, the results showed that the order of presentation of the clinical vignettes could affect participants’ responses, suggesting that future iterations of the tool may benefit from refining or standardizing the vignette sequence to reduce this bias. Also, in this project, we were not able to analyze test-retest reliability, which involves repeated administration of the instrument to the same participants on several occasions, in order to determine the degree to which the participant’s performance is repeatable and to assess the internal consistency of the instrument. As the completion of the instrument is based on a luring strategy (i.e. the real purpose of the survey is concealed from participants until the end of the questionnaire), this step could not be carried out, as it would have introduced a bias in responses in the second administration.

Finally, although the tool includes several dimensions of stigmatization, it could benefit from further development to capture more specific aspects of stigmatization that might arise in specialized clinical contexts. These limitations highlight the need for additional research and testing to ensure the instrument’s robustness and broader applicability across healthcare professions and settings.

## 6. Conclusion

This participatory work has led to the development of a highly innovative tool for assessing a crucial social issue: the stigmatization of Indigenous patients in clinical contexts. Thanks to its original format, it is one of the only tools of its kind in the Western context. Although the tool still needs to be refined, it has shown definite psychometric qualities. Further tests and adaptations will allow to make it more robust.

## Data Availability

The minimal data set will be made available on a institutional repository.

## Acknowledgement

The team would like to acknowledge France Robertson, director of the Lanaudière Indigenous Friendship Centre who passed away in 2018, with whom the initial discussions about this project took place. The team would also like to acknowledge the work and contribution of Élaine Brière, a patient partner from the Wolastoqiyik Wahsipekuk nation who passed away in 2026. Élaine was part of the research team from the very beginning and provided constant ideas and feedback throughout all phases of the tool’s development.

